# Base Editing Gene Therapy for Heterozygous Familial Hypercholesterolemia

**DOI:** 10.1101/2025.04.17.25325983

**Authors:** Ping Wan, Siyuan Tang, Dongni Lin, Yuming Lu, Mei Long, Ling Xiao, Yanhong Jiang, Jiaoyang Liao, Xiaoying Ma, Ying Liu, Wensu Yu, Zi Jun Wang, Yuxuan Wu, Taihua Yang, Qiang Xia

**Author notes:** Drs. Wan, Tang, Lin, Lu and Long contributed equally to this article.

## Abstract

**Background:** Heterozygous familial hypercholesterolemia (HeFH) is a genetic disorder characterized by persistently elevated low-density lipoprotein cholesterol (LDL-C) levels, leading to an increased risk of early-onset atherosclerosis cardiovascular diseases (ASCVD). YOLT-101, an in vivo base-editing therapeutic agent delivered via GalNAc-modified lipid nanoparticles, is designed to achieve permanent inactivation of proprotein convertase subtilisin/kexin type 9 (*PCSK9*), enabling sustained LDL-C reduction.

**Methods:** This trial enrolled participants with heterozygous genetic mutations in the low-density lipoprotein receptor (LDLR), and LDL-C levels of ≥2.6 mmol/L (without ASCVD) or ≥1.8 mmol/L (with ASCVD) despite receiving moderate- or high-intensity statin therapy. Eligible patients received a single intravenous infusion of YOLT-101 at ascending doses (0.2, 0.4, and 0.6 mg/kg). We report interim results from an ongoing clinical trial evaluating the safety, tolerability, pharmacodynamics, and efficacy of YOLT-101.

**Results:** Six participants were enrolled (median age, 48 years, range, 34-62) in the study. The most common adverse events (AEs) were transient infusion-related reactions (83.3%) and elevations in alanine/aspartate aminotransferase (50%). No study withdrawals or AEs of grade 3 or higher occurred. PCSK9 and LDL-C levels decreased in a dose-dependent manner following YOLT-101 administration. In the 0.6mg/kg group (n=3), mean PCSK9 levels decreased by 55.9% at week 1 and by 75.8% and 72.5% after 1 and 4 months, respectively; corresponding LDL-C reductions were 33.2%, 48.9%, and 50.4%, respectively.

**Conclusions:** A single infusion of YOLT-101 at 0.6 mg/kg was well tolerated and led to sustained PCSK9 and LDL-C reduction, demonstrating promise for future clinical development. (Funded by YolTech Therapeutics; Registration Number: NCT06458010)

## INTRODUCTION

Heterozygous familial hypercholesterolemia (HeFH) is a prevalent autosomal dominant genetic disorder, affecting an estimated 1 in 250 to 1 in 300 individuals globally^1,2^. This condition is defined by defective metabolism of low-density lipoprotein cholesterol (LDL-C), leading to persistently elevated plasma LDL-C levels from birth. If left untreated, HeFH confers a markedly increased risk of early-onset atherosclerotic cardiovascular disease (ASCVD), which poses a major threat to public health due to the substantial patient population^3^. The molecular etiology of HeFH is predominantly attributed to mutations in genes critical to LDL-C regulation, with over 90% of cases involving the low-density lipoprotein receptor (*LDLR*), approximately 5% involving the apolipoprotein B (*APOB*)^1^, and fewer than 2% involving the proprotein convertase subtilisin/kexin type 9 (*PCSK9*)^4^. The therapeutic goal of HeFH is the long-term control of elevated serum LDL-C levels through lifestyle management and cholesterol-lowering therapy based on patients’ risks and severity of ASCVD to minimize cumulative cholesterol burden^5^.

First-line lipid-lowering drugs include high-intensity statins, such as atorvastatin, rosuvastatin or pitavastatin^6,7^. Ezetimibe, bempedoic acid, and other adjunctive drugs are often added if LDL-C targets are not met^8^. However, a subset of patients exhibit suboptimal responses to lipid-lowering drugs, even at maximal tolerated doses^9–11^. PCSK9-targeted therapies have emerged as effective treatments^12^, reducing LDL-C either by directly inhibiting *PCSK9* function or suppressing its expression, thereby preventing the lysosomal degradation of LDLRs, with no apparent adverse effects^13^. Monoclonal antibodies such as alirocumab and evolocumab neutralizing circulating PCSK9 were initially introduced in treatments for HeFH patients, and administered biweekly or monthly^14,15^. More recently, the second-generation PCSK9 inhibitor, exemplified by inclisiran—a small interfering RNA (siRNA) agent—has demonstrated a more persistent LDL-C reduction with biannual administration by silencing hepatic *PCSK9* expression^16^. The effectiveness of these PCSK9-directed therapies relies on strict adherence to lifelong treatment, which is challenging for a considerable number of patients due to difficulties with self-injection, limited awareness of cardiovascular risks, or high medication costs. Discontinuation, however, leads to a gradual return of PCSK9 and LDL-C levels to baseline. Furthermore, even with the combined use of maximal tolerated doses of statins and inclisiran (with or without ezetimibe), over 30% of HeFH patients failed to achieve the LDL-C goal levels, remaining exposed to the risk of cardiovascular disease progression^16^. Given these limitations, there is a compelling need for more effective and durable treatments for HeFH.

Gene editing technologies have advanced rapidly, transitioning from basic research to practical clinical applications. The approval of Casgevy, the first CRISPR/Cas9-based therapy, marked a milestone in ex vivo gene editing. This therapy restored fetal hemoglobin (HbF) expression in CD34[ hematopoietic stem/progenitor cells from patients with β-hemoglobinopathies by editing the BCL11A enhancer, resulting in significant clinical benefits^17,18^. In the NTLA-2001 trial, lipid nanoparticles (LNPs) were utilized to deliver the CRISPR/Cas9 system directly to hepatocytes, successfully achieving targeted gene knockout^19^, marking the beginning of the in vivo CRISPR/Cas9 era. While CRISPR/Cas9 editing relies on double-strand breaks (DSBs)^20–22^, base editing enables direct nucleotide conversion without introducing DSBs, thereby reducing off-target effects and enhancing precision^23^. In a previous study, we developed a high-precision adenine base editor (hpABE5) derived from a naturally occurring adenine deaminase. Using AlphaFold-driven structural predictions and analyses of 759 enzyme candidates, we selected TadA from Hafnia paralvei (hpTadA) and enhanced its deaminase activity and precision through iterative saturation mutagenesis. The resulting hpABE5 exhibits high editing efficiency, a refined editing window, and minimized off-target effects. YOLT-101, an in vivo gene-editing drug administered via intravenous infusion, utilizes guide RNA (gRNA) and hpABE5 mRNA to edit *PCSK9* for the lowering of LDL-C. Upon translation, the ABE protein forms a complex with the gRNA to target the *PCSK9* gene, introducing an A-to-G substitution without DSBs. This precise editing disrupts PCSK9 mRNA splicing, introduces a frameshift mutation, and effectively silences *PCSK9* expression, thereby reducing LDLR degradation and lowering LDL-C levels^20^ (Fig. 1). The drug employs a novel LNP to deliver the RNAs to hepatocytes. Instead of solely utilizing the LDLR pathway on the surface of hepatocytes, which is partially defective in HeFH patients with LDLR mutants, we utilized an optimized LNP capable of delivering the editor via LDLR-independent pathways, with additional targeting of the asialoglycoprotein receptor (ASGPR) via N-acetylgalactosamine (GalNAc)^24^. Altogether, YOLT-101 offers the potential for single-dose, DNA-level interventions capable of conferring lifelong therapeutic benefits for patients with HeFH.

**Figure 1.**
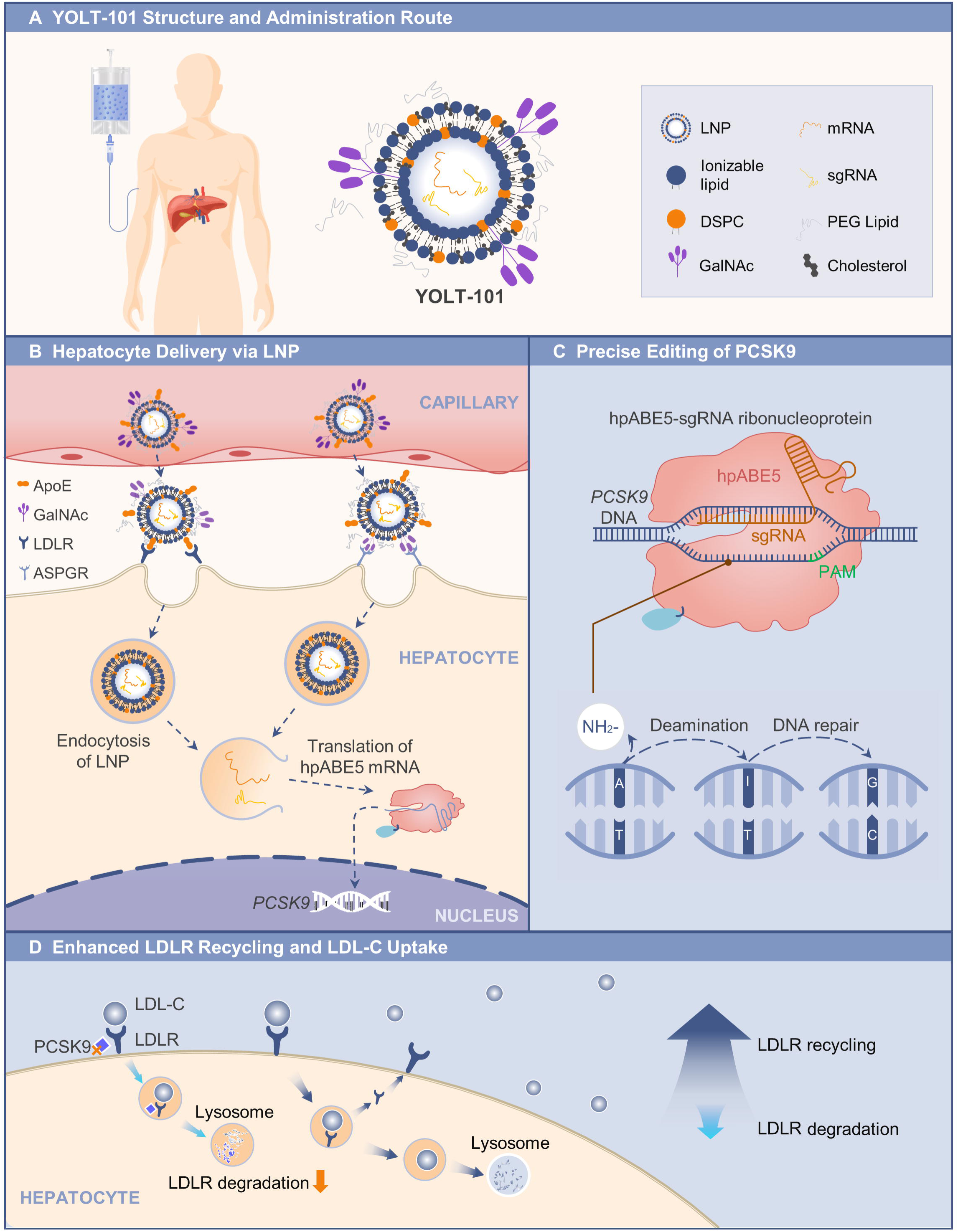
Mechanism of YOLT-101 in Reducing PCSK9 and Lowering LDL-C. Panel. **A** shows the structure of YOLT-101, which is administered via intravenous infusion. Its carrier system is a GalNAc-modified lipid nanoparticle (LNP) system, designed for enhanced targeted delivery to hepatocytes. The active components, including adenine base editor (hpABE5) mRNA and single-guide RNA (sgRNA), are encapsulated within the LNP and specifically target the proprotein convertase subtilisin/kexin type 9 (*PCSK9*) gene in the liver. **Panel B** illustrates the dual-targeting mechanism by which YOLT-101 is delivered to hepatocytes. Following administration and entry into the circulation, the LNPs are opsonized by apolipoprotein E (ApoE). The LNPs, which contain the active components, bind to the low-density lipoprotein receptor (LDLR) on hepatocyte surfaces via ApoE, facilitating receptor-mediated endocytosis. Simultaneously, the LNPs are directed to hepatocytes through GalNAc, which specifically targets the asialoglycoprotein receptor (ASGPR) on hepatocyte surfaces, enhancing delivery via an LDLR-independent pathway. **Panel C** shows the precise base editing process. Upon entering the hepatocyte, sgRNA guides the hpABE5 ribonucleoprotein complex to the target site in the *PCSK9* gene. The complex catalyzes the deamination of adenine (A) to inosine (I), which is then processed by the cell’s DNA repair machinery as guanine (G). This results in a precise A-to-G substitution, disrupting the normal splicing of PCSK9 mRNA and introducing a frameshift mutation that inactivates the *PCSK9* gene, effectively silencing its expression. **Panel D** illustrates that the reduction in *PCSK9* expression leads to enhanced recycling of the LDLR. With less PCSK9 available to degrade LDLR, the receptors remain functional and are efficiently recycled, increasing LDL-C uptake into hepatocytes. This process lowers circulating LDL-C levels, thereby resulting in the therapeutic effect of YOLT-101.

In this study, we present interim findings from an ongoing clinical trial investigating the safety, tolerability, and efficacy of single ascending doses of YOLT-101 for *PCSK9* gene editing in HeFH patients. These data highlight the transformative potential of our base editing therapy in achieving sustained LDL-C reduction in this population at high risk of ASCVD.

## METHOD

### Preclinical Analysis of Off-Target Effects

We selected a single-guide RNA (sgRNA) targeting the exon 1 splice-donor region of PCSK9 (CCCGCACCUUGGCGCAGCGG; hg38, chr1: 55040032–55040051) to induce adenine-to-guanine conversion using the high-precision adenine base editor hpABE5 (PCT/CN2023/078133; PCT/CN2024/127500; CN202410212048.2), with editing specificity confirmed by extensive off-target analysis. Potential gDNA-dependent off-target loci were identified using Digenome-seq, CIRCLE-seq, and cellular-based GUIDE-seq. Off-target effects were evaluated in primary human hepatocytes at the EC90 dose, with sequencing depth exceeding 20,000x to ensure high sensitivity. The gDNA-independent off-target editing was assessed by whole-genome sequencing (WGS), while RNA-dependent off-target effects were analyzed through RNA sequencing (RNA-seq). Chromosomal translocations and other abnormalities at the on-target site were examined using PacBio sequencing. A PacBio library was constructed with a hybrid capture method to sequence approximately a 25 kbp region surrounding the on-target site at an exceptionally high depth of over 25,000x.

### Trial Design and Oversight

This clinical trial (NCT06458010) was designed as a single-arm, open-label, single-dose escalation study, initialized by investigators in Ren Ji Hospital, Shanghai Jiao Tong University School of Medicine in Shanghai, China. The trial protocol was developed collaboratively by the investigators of Ren Ji Hospital and medical experts from YolTech Therapeutics. It was reviewed by national and institutional ethic and regulatory bodies, including expert committees responsible for the assessment of new studies of gene therapy. All patients underwent screening, enrollment and treatment at Ren Ji Hospital Clinical Trial Centre. The study was strictly implemented under the Declaration of Helsinki. Written informed consent was obtained from all participants prior to enrollment. Data collection, analysis, and interpretation were conducted by the research team, with all authors contributing to manuscript preparation and the decision to submit it for publication. The authors affirm the accuracy and completeness of the data and confirm that the trial was conducted in strict adherence with the protocol.

### Patients

Eligibility criteria for enrollment included a confirmed diagnosis of HeFH through genetic testing. Participants were required to have LDL-C levels of at least 100 mg/dL (2.6 mmol/L) despite receiving moderate- or high-intensity statin therapy (maintained on a stable regimen for ≥4 weeks) or ≥70 mg/dL (1.8 mmol/L) for those with a history of ASCVD. Patients were excluded if they had received PCSK9 monoclonal antibody therapy within 3 months or PCSK9 small nucleic acid-based therapies within 1 year prior to screening. Additionally, individuals participating in other lipid-lowering clinical trials or using investigational lipid-lowering agents were excluded.

### Trial Procedures

Participants received a single intravenous infusion of YOLT-101 at doses of 0.2 mg/kg (n=1), 0.4 mg/kg (n=2), or 0.6 mg/kg (n=3), administered for more than 2 hours. To mitigate infusion-related reactions, patients were pretreated with 7.5 mg of oral dexamethasone (or equivalent) 8-24 hours prior to infusion, followed by a single dose of a steroid (dexamethasone 10 mg, intravenously), histamine H1 receptor antagonist (levocetirizine 10 mg, orally), and histamine H2 receptor antagonist (famotidine 20 mg, orally) 30-60 minutes before YOLT-101 administration. Patients were required to fast for at least 9 hours before the infusion. All participants will be monitored in the hospital for at least 7 days following the treatment. The follow-up consists of the main study follow-up within the first year, as well as long-term follow-up from the 2nd to the 15th year. We report interim results from the ongoing clinical trial.

### Clinical Assessments and End Points

The study was designed to evaluate the safety, tolerability, pharmacodynamics and efficacy profiles of a single administration of YOLT-101 in patients with HeFH and also to identify the optimal biologically active dose (OBD). These evaluations were conducted using standardized protocols. Safety and tolerability evaluations included monitoring adverse events (AEs; graded according to Common Terminology Criteria for Adverse Events v5.0), vital signs, physical examinations, 12-lead electrocardiograms (ECGs), complete blood counts, liver and renal function tests, coagulation profile assessments, immunogenicity tests, and measurements of the C-reactive protein, complements, cytokines, and lymphocyte subsets. Pharmacodynamic effects were assessed by measuring the percentage change in plasma PCSK9 levels at predefined time points relative to baseline using enzyme-linked immunosorbent assay (ELISA). Efficacy was evaluated based on changes in plasma LDL-C levels, determined using the Cobas system, which employs electrochemiluminescence technology, at the corresponding post-administration time points.

### Statistical Analysis

All data were presented descriptively. Safety and tolerability assessments involved data from all enrolled patients. Absolute and percent changes of plasma PCSK9 and LDL-C from baseline levels were analyzed using data from patients who had been maintained on a fixed lipid-lowering regimen since at least 28 days before YOLT-101 therapy.

## RESULTS

### Preclinical Off-Target Data

The preclinical development of YOLT-101 included computational modeling and in vitro off-target studies to ensure precise on-target gene editing. In primary human hepatocytes, YOLT-101 was highly potent (EC90, 2-2.5 ng/mL) and achieved efficient *PCSK9* editing.

To rigorously assess gDNA-dependent off-target effects, potential off-target loci were identified using Digenome-seq, CIRCLE-seq, and GUIDE-seq. A total of 62 potential off-target sites were detected, with 9 confirmed by at least two methods and 1 identified by all three approaches. Targeted sequencing of these sites revealed no detectable off-target editing when primary human hepatocytes were treated with YOLT-101 at the EC90 dose. Additionally, the gDNA-independent off-target editing at the DNA level was evaluated by WGS. At the EC90 dose, no significant difference in A to G or T to C mutations was observed between the treated and untreated primary human hepatocytes.

Adenine base editors have been shown to induce gRNA-independent RNA editing via the deoxyadenosine deaminase domain^25^. To assess RNA editing, RNA sequencing (RNA-seq) was performed on primary human hepatocytes treated with YOLT-101. No significant additional A-to-G RNA edits were detected in treated hepatocytes compared to controls.

To evaluate potential genomic alterations, long-read sequencing using Pacific Biosciences technology was performed at a saturating dose to detect insertions, duplications, and large deletions. No abnormal genomic changes were observed compared to controls. These findings support the specificity and safety profile of YOLT-101 in primary human hepatocytes, demonstrating efficient *PCSK9* editing with minimal off-target effects.

### Participants

Eight patients completed the screening assessment, of whom six were ultimately enrolled in the dose-escalation trial. Two patients were excluded due to unstable angina and a plan for coronary stenting respectively. The overall cohort demonstrated a median age of 48 (range, 34–62) years and a median body weight of 74.3 (range, 51.5–79.5) kg, with an equal gender distribution (3 females and 3 males). Genetic analysis revealed a diverse spectrum of LDLR mutations among participants. The most frequently observed mutation, c.1448G>A, was identified in one patient in each of the 0.4 mg/kg and 0.6 mg/kg groups. The remaining four patients carried distinct mutations: c.817+1G>C, c.973T>C, c.817+1_817+50del, and c.1476_1477CT. In terms of cardiovascular assessment, five (5/6, 83.3%) patients demonstrated evidence of coronary artery stenosis as detected by coronary computed tomography angiography (CCTA), with the left anterior descending artery (LAD) and right coronary artery (RCA) representing the most frequently implicated vascular segments, each observed in 50% of the cases. Furthermore, ASCVD was diagnosed in two patients (2/6, 33.3%), with one individual belonging to the 0.2 mg/kg dosing cohort and the other to the 0.4 mg/kg cohort.

Patients 1 and 2, who had not received lipid-lowering therapy within 28 days prior to YOLT-101 administration, were started on lipid-lowering therapy 28 days and 2 days after YOLT-101 administration, respectively, to mitigate potential cardiovascular risks. Patients 3-6 have been maintained on a fixed regimen of rosuvastatin (10 mg/day) at least 28 days before YOLT-101 therapy to evaluate the independent effects of YOLT-101 on lipid profiles. The mean (±SD) PCSK9 and LDL-C values of the 6 patients before YOLT-101 treatments were 598.7±126.7 μg/L and 186.9±22.0 mg/dL (4.82±0.57 mmol/L), respectively. Additional baseline characteristics are detailed in Table 1.

**Table 1.**
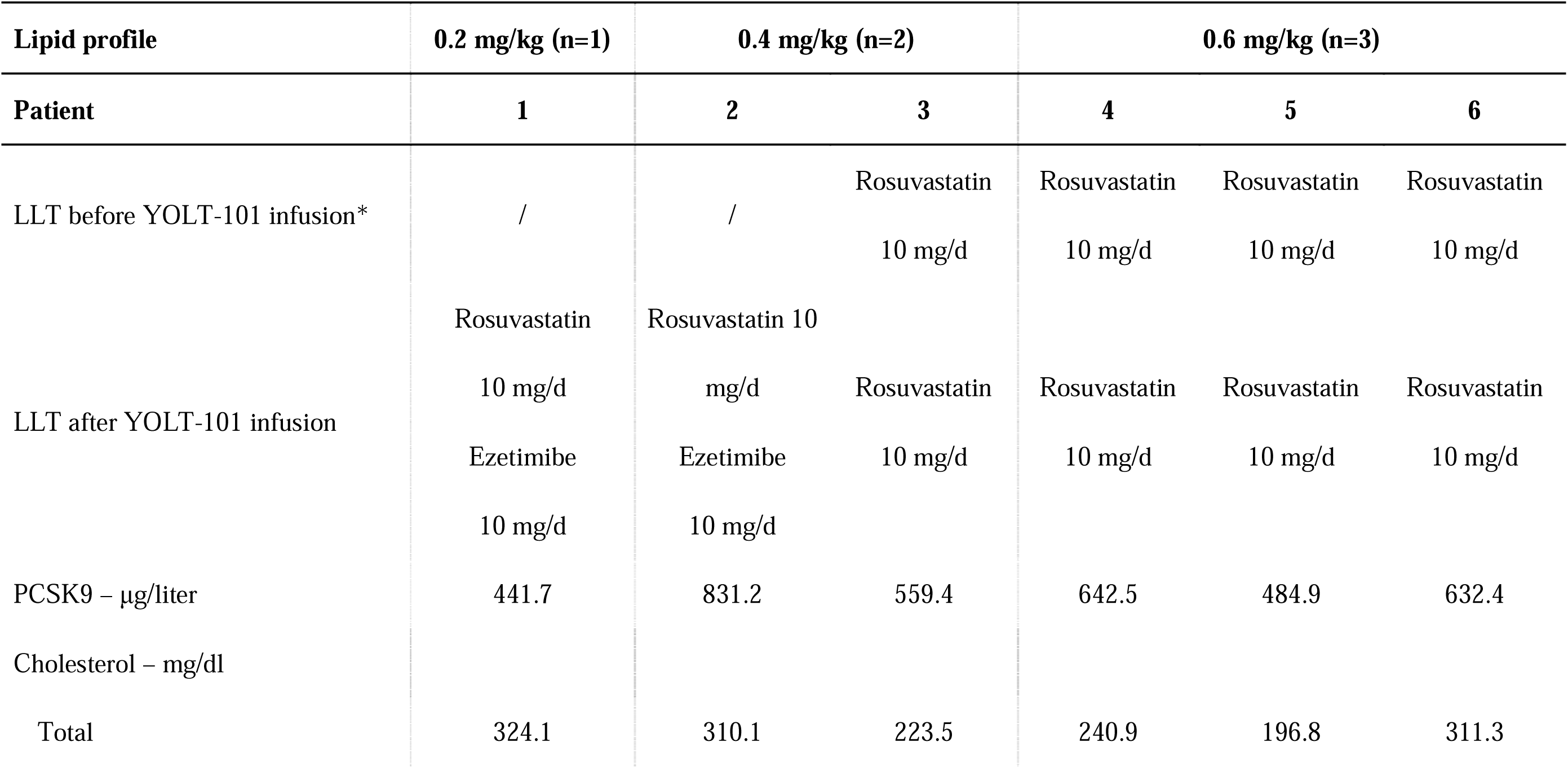
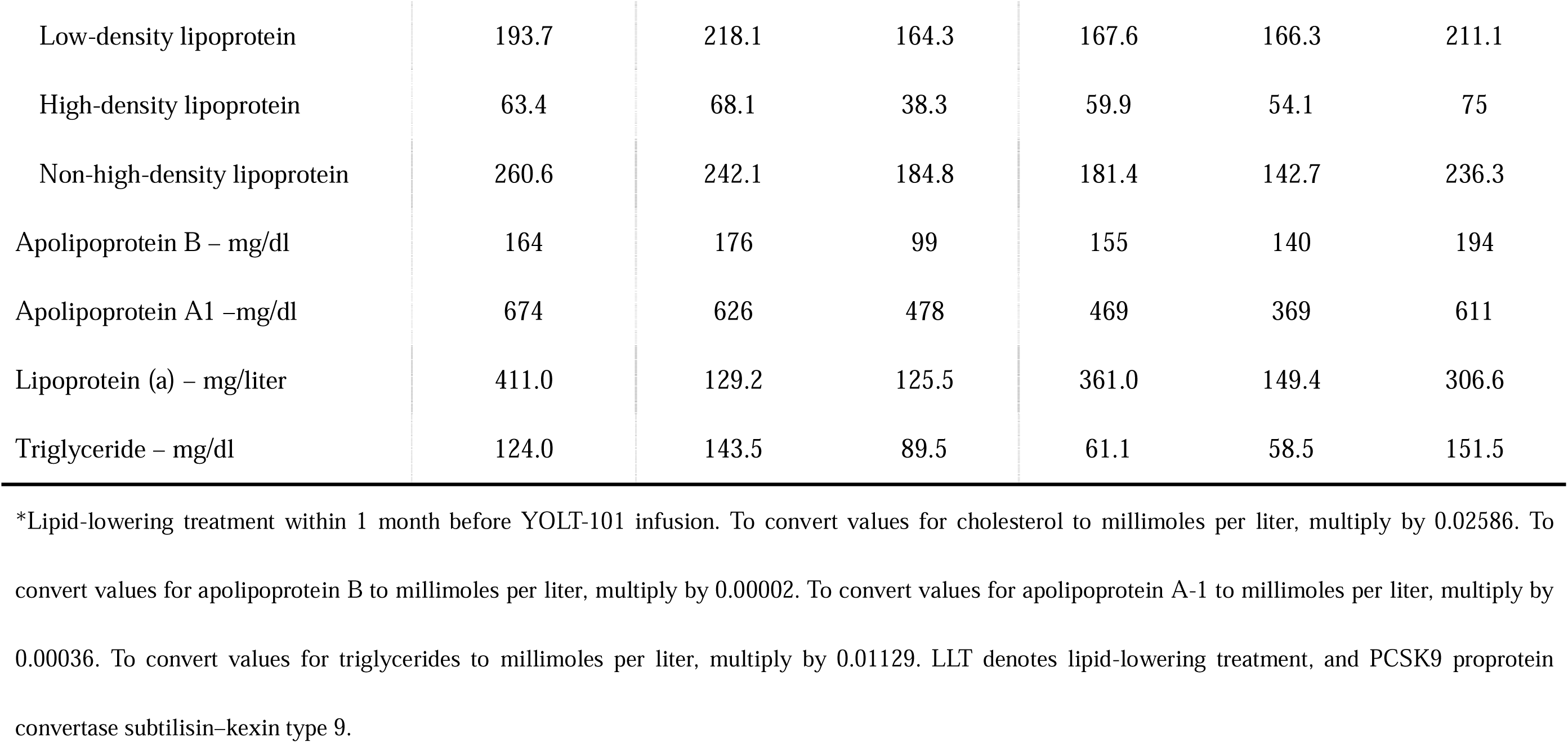
Baseline Lipid Profiles and Lipid-lowering Treatments of Patients.

### Safety and Side-Effect Profile

The safety profile of YOLT-101 was assessed in six patients across three dose cohorts: 0.2 mg/kg (n=1), 0.4 mg/kg (n=2), and 0.6 mg/kg (n=3). As of January 31, 2025, the median follow-up duration was 6.5 (range, 4-9) months. Treatment-related AEs were reported in five out of six patients (83.3%), all of which were mild to moderate in severity. The most frequently observed treatment-related adverse events were the infusion-related reactions (including symptoms such as fever, myalgia, and vomiting) which occurred in all five patients (5/6, 83.3%) receiving YOLT-101 at doses of 0.4 mg/kg or 0.6 mg/kg. Infusion-related reactions typically occurred within 8-10 hours following YOLT-101 treatments and recovered within 24 hours. One patient (1/6, 16.7%) in the 0.4 mg/kg cohort, who had a history of ASCVD, experienced chest pain approximately 6 hours after administration of YOLT-101, but symptoms improved with oxygen supplementation (Table 2). Transient elevations in alanine aminotransferase (ALT) and aspartate aminotransferase (AST) were noted in 3 patients (50.0%), without any elevations in total bilirubin (TBIL). ALT and AST levels almost returned to the normal range within one month. A treatment-unrelated adverse event, nausea, was found in one patient in the 0.4 mg/kg cohort (1/6, 16.7%). No serious adverse events (SAEs), AEs of grade 3 or higher as per the Common Terminology Criteria for Adverse Events (CTCAE) version 5.0, or any events leading to discontinuation of YOLT-101 or withdrawal from the trial were reported. There were no deaths during the study period (Table 2).

**Table 2.** Adverse Events and Key Safety Laboratory Results*.

| **Event** | **0.2 mg/kg (n=1)** | **0.4 mg/kg (n=2)** | **0.6 mg/kg (n=3)** | **Overall (n=6)** |
| --- | --- | --- | --- | --- |
|  | **no. (%)** | **no. (%)** | **no. (%)** | **no. (%)** |
| At least one adverse event | 0 | 2 (100.0) | 3 (100.0) | 5 (83.3) |
| Treatment-related adverse event |  |  |  |  |
| Infusion-related reaction | 0 | 2 (100.0) | 3 (100.0) | 5 (83.3) |
| Alanine aminotransferase elevation | 0 | 1 (50.0) | 2 (66.6) | 3 (50.0) |
| Aspartate aminotransferase elevation | 0 | 1 (50.0) | 2 (66.6) | 3 (50.0) |
| Chest pain | 0 | 1 (50.0) | 0 | 1 (16.7) |
| Treatment-unrelated adverse event |  |  |  |  |
| Nausea | 0 | 1 (50.0) | 0 | 1 (16.7) |
| Any serious event | 0 | 0 | 0 | 0 |
| Any adverse event ≥ grade 3 in CTCAE severity | 0 | 0 | 0 | 0 |
| Adverse event leading to discontinuation of YOLT-101 | 0 | 0 | 0 | 0 |
| Adverse event leading to withdrawal from the trial | 0 | 0 | 0 | 0 |
| Death | 0 | 0 | 0 | 0 |

\*Severity of all adverse events was assessed via the National Cancer Institute Common Terminology Criteria for Adverse Events (CTCAE) version 5.0.

### PCSK9 levels

PCSK9 levels were evaluated at multiple time points. Data from patients 3 through 6, who maintained consistent lipid-lowering drug regimens throughout the study period, were analyzed to assess the independent effects of YOLT-101 on PCSK9 levels. In the 0.4 mg/kg cohort, an 8.6% decrease in PCSK9 levels was observed one week after YOLT-101 administration. The decrease persisted, with a 15.8% reduction at two weeks and a 30.4% reduction at four weeks following treatment. In the 0.6 mg/kg cohort, the mean (±SD) PCSK9 levels decreased by 55.9±25.7% one week after YOLT-101 therapy compared to baseline values, followed by a further decline to a 64.2±9.0% reduction at two weeks and 75.8±7.1% at four weeks (Fig. 2A and 2B). Plasma PCSK9 levels gradually stabilized and reached a plateau after four weeks, maintaining a consistently low level. At four weeks, the absolute values had dropped from 559.4 μg/L to 389.6 μg/L in the 0.4 mg/kg group, and the mean (±SD) value in the 0.6 mg/kg group dropped from 586.6±72.0 μg/L to 147.3±55.3 μg/L (Fig. 2C). The 2 dosage groups exhibited a clear dose-dependent response, with higher doses correlating with more substantial reductions in PCSK9 levels.

**Figure 2.**
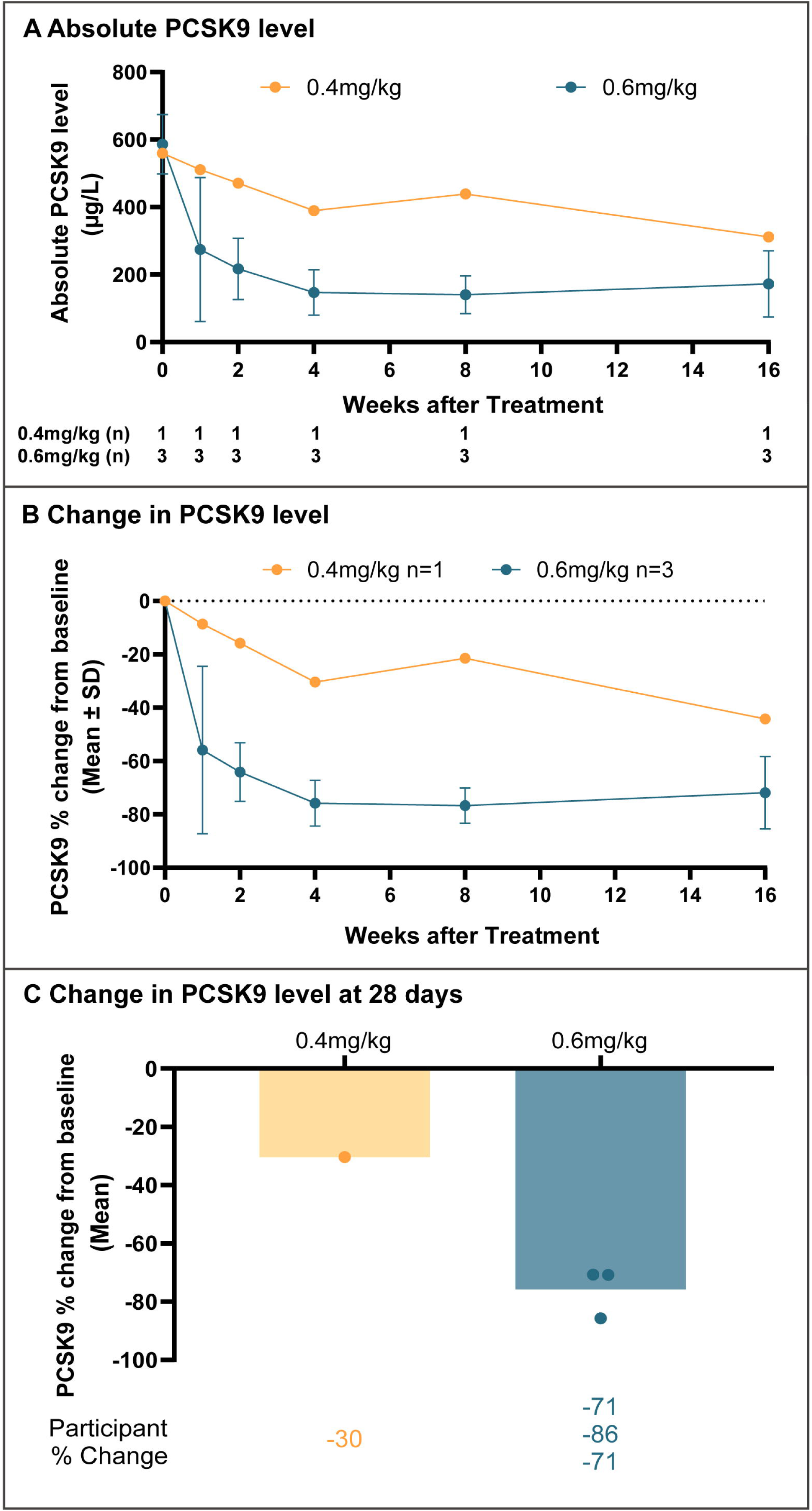
Changes in PCSK9 Levels Following YOLT-101 Administration. Panel A shows absolute PCSK9 levels over time in patients 3 through 6 treated with YOLT-101. Panel B shows percentage changes in PCSK9 levels over time from baseline in patients 3 through 6 treated with YOLT-101. Panel C shows percentage changes in PCSK9 levels at 28 days post-treatment, stratified by dosing cohorts (0.4 mg/kg and 0.6 mg/kg). Data are presented as mean `±` standard deviation (SD) or individual patient values, as indicated.

### LDL-C Levels

LDL-C levels were also evaluated using data from patients 3 through 6. In the 0.4 mg/kg cohort, LDL-C levels began to decrease by 12.9% seven days post-administration, with further reductions of 20.2% and 33.9% observed at the two- and four-week time points, respectively. In the 0.6 mg/kg cohort, a pronounced 33.2±14.6% mean (±SD) reduction in LDL-C levels was observed one week after YOLT-101 therapy, decreasing to 123.9±40.0 mg/dL (3.20±1.03 mmol/L) from a baseline value of 181.7±20.8 mg/dL (4.70±0.54 mmol/L). At 2 and 4 weeks following treatments, the mean reductions of 38.1±18.1% and 48.9±18.4% were recorded compared to the baseline level. Thereafter, LDL-C levels gradually plateaued. Notably, at four weeks post-YOLT-101 administrations, the mean (±SD) LDL-C levels in the 0.4 mg/kg and 0.6 mg/kg cohorts declined to 108.7 mg/dL (2.81 mmol/L) and 94.2±36.8 mg/dL (2.44±0.95 mmol/L), respectively, highlighting a dose-dependent lipid-lowering efficacy of YOLT-101 (Fig. 3A and 3B). These findings suggested that the reduction in PCSK9 levels was accompanied by a significant decrease in LDL-C levels.

**Figure 3.**
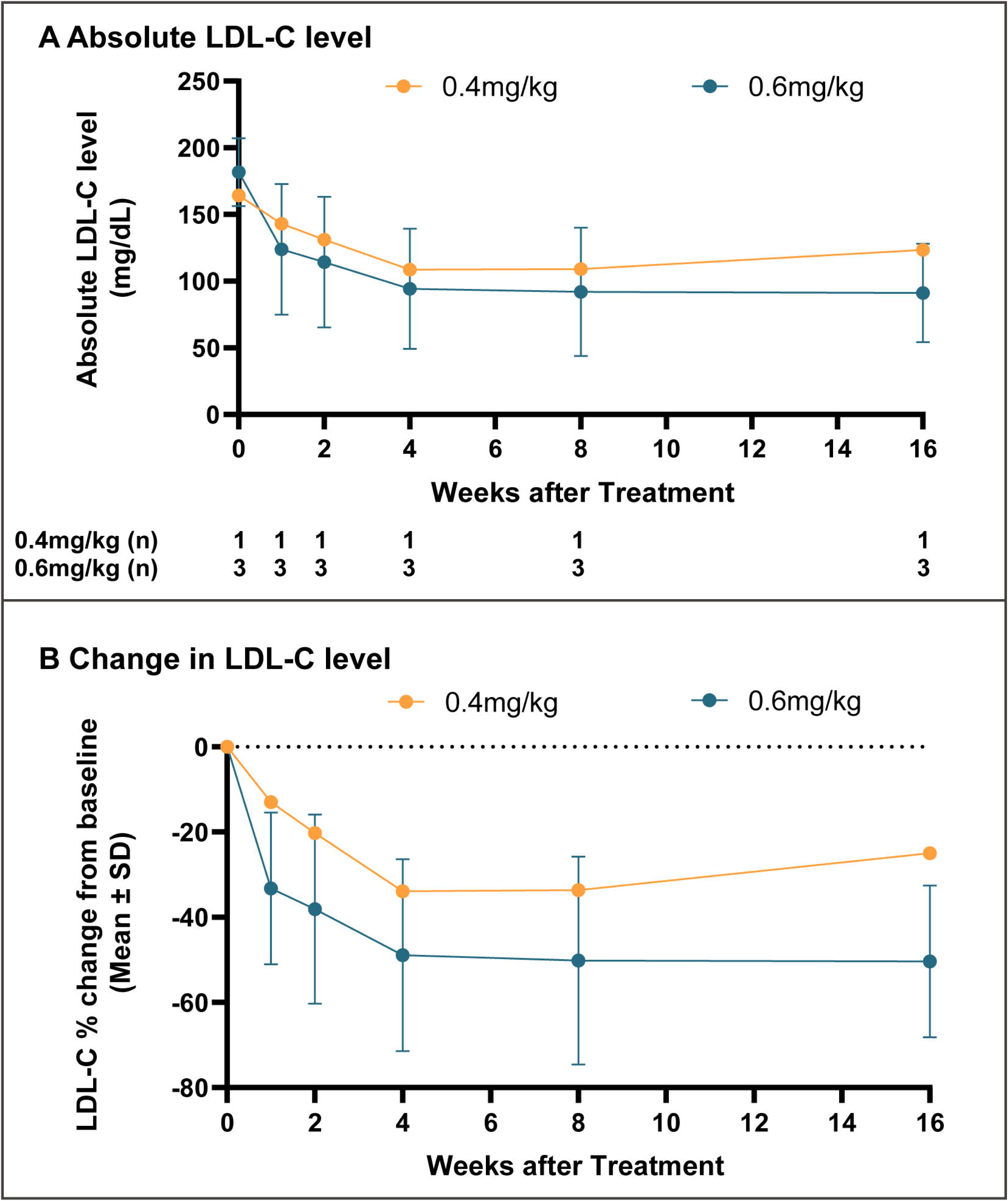
Changes in LDL-C Levels Following YOLT-101 Administration. Panel A shows absolute LDL-C levels over time in patients 3 through 6 treated with YOLT-101. Panel B shows percentage changes in LDL-C levels over time from baseline in patients 3 through 6 treated with YOLT-101. Data are presented as mean ± standard deviation (SD), as indicated. To convert values for cholesterol to millimoles per liter, multiply by 0.02586.

## DISCUSSION

Patients with HeFH require lifelong lipid-lowering therapy to mitigate the risk of ASCVD. The International Atherosclerosis Society guidelines recommend LDL-C targets for HeFH: <100 mg/dL (2.6 mmol/L) without ASCVD or major risk factors, <70 mg/dL (1.8 mmol/L) with imaging evidence of ASCVD or major risk factors, and <55 mg/dL (1.4 mmol/L) for clinical ASCVD patients^5,26^. Inclisiran, a small interfering RNA targeting hepatic PCSK9 synthesis, provides a new treatment option for HeFH patients who may not respond well to traditional statin therapy or those who cannot tolerate statins due to side effects. For HeFH patients already receiving the maximum dose of statins with or without ezetimibe, the incorporation of inclisiran administered twice yearly (after initial and 3-month doses), further decreased the LDL-C level by 38.1%. However, 30.2% of patients still did not achieve the recommended target LDL-C goals despite the addition of inclisiran, remaining exposed to the risk of cardiovascular disease progression^16^. Furthermore, the requirement for repeated administration throughout patients’ lifetime also presents a limitation^27,28^. Poor adherence to the long-term lipid-lowering therapy, often due to side effects of drugs, high medication fees, and unawareness of the disease severity, posed a significant challenge for a substantial number of patients^29^. Recent advances in CRISPR/Cas9-based gene therapy and associated delivery technologies presented the potential for a one-time, durable treatment option, addressing the unmet need for sustained lipid control while enhancing patient convenience and adherence.

In this study, we present the first-in-human evidence of YOLT-101, an in vivo ABE therapy for HeFH patients. Six HeFH patients with LDLR mutation received ascending doses of YOLT-101 (0.2, 0.4, or 0.6 mg/kg). A 72.5% reduction in PCSK9 levels was recorded on average in patients receiving YOLT-101 at a dose of 0.6 mg/kg, sixteen weeks after treatment, accompanied by an average 50.4% reduction in LDL-C compared to baseline levels. In contrast, only a 60.7% reduction in PCSK9 and a 38.1% reduction in LDL-C were observed in HeFH patients receiving PCSK9-targeted siRNA drug, inclisiran. Thus, a single dose of YOLT-101 showed more pronounced decreases in both PCSK9 and LDL-C levels. Moreover, for patients who might not have reached the target LDL-C level after the initial YOLT-101 treatment, a second dose provides an opportunity for complete PCSK9 gene editing in the liver, thereby further enhancing therapeutic efficacy. All AEs were mild to moderate in severity, with transient infusion-related reactions and reversible liver enzyme elevations (ALT/AST) being the two most common AEs observed. No serious AEs, dose-limiting toxicities, or treatment discontinuations occurred. Thus, YOLT-101 demonstrates both efficacy and convenience advantages, offering the potential for a durable, one-time alternative to continuous inclisiran injections, while maintaining a favorable safety profile.

Recently, Gillmore JD and colleagues have published data on patients with transthyretin amyloidosis using CRISPR-Cas9 in vivo gene editing and LNP delivery system to target and disrupt the *TTR* gene, thereby reducing the production of misfolded transthyretin protein and mitigating disease progression^19^. CRISPR-Cas9 gene editing has been increasingly utilized as a powerful tool for precise genetic modifications, offering potential therapeutic solutions for various genetic disorders^30^. The implementation of CRISPR-Cas9 in biomedical research and clinical applications has paved the way for innovative treatments, enabling long-term and potentially curative interventions. However, this technology relies on DSBs to introduce edits, which can lead to indels or insertions and carry risks of genomic instability and off-target effects, as well as chromosomal rearrangements and p53-mediated DNA damage responses^31–33^. By contrast, YOLT-101 employs the LNP conjugated with GalNAc to carry an ABE system without creating DSBs, making it a potentially safer and more accurate option.

Furthermore, we employed a GalNAc-modified LNP as the delivery vehicle, specifically designed to address the challenges posed by reduced LDLR function in liver-targeted drug delivery for patients with HeFH. Typically, LNPs rely on the adsorption of apolipoprotein E (APOE) and subsequent uptake via the LDLR pathway to deliver their payload into hepatocytes^34^. However, in HeFH patients, the reduced number of LDLRs impairs LNP uptake efficiency, limiting the liver delivery of drugs. To overcome this challenge, we engineered LNPs with GalNAc, a ligand that binds to ASGPR, which is highly expressed in hepatocytes^35^. This modification enables the therapeutic payload to be delivered to hepatocytes without relying entirely on LDLR-mediated uptake, thereby improving cellular uptake and ensuring robust therapeutic effects^24^. This delivery strategy not only overcomes the limitations imposed by LDLR deficiency in HeFH patients but also highlights the potential of receptor-targeted nanomedicine for treating other genetic disorders associated with impaired receptor functions.

Given the therapeutic implications, it was essential to thoroughly assess the safety and specificity of YOLT-101, particularly in its potential for off-target gene editing. To this end, a comprehensive analysis was conducted, focusing on three key areas: the identification and validation of potential gDNA-dependent off-target sites, the evaluation of gDNA-independent off-target effects, and an analysis of genome stability. Computational modeling, biochemical cell-free assays, and in vitro cellular assays were employed to predict and identify potential off-target sites outside the *PCSK9* gene. Importantly, no evidence of gDNA-dependent off-target events, gDNA-independent off-target events, or chromosomal abnormalities was detected. Additionally, participants receiving YOLT-101 therapy will require long-term safety monitoring.

This trial remains ongoing, with larger patient cohorts to further validate the therapeutic durability and safety profile. Preliminary data from the initial cohort demonstrate clinical validation of adenine base editing for downregulating PCSK9 and lowering LDL-C, achieving sustained *PCSK9* editing without detectable off-target activity, thereby supporting single-dose genome editing as a therapeutic strategy for HeFH.

## Data Availability

All data produced in the present study are available upon reasonable request to the authors

